# Loss of H3K9 di-methylation perpetuates the type I interferon signature in SLE and is pharmacologically reversible by ribavirin

**DOI:** 10.1101/2025.08.04.25332258

**Authors:** Qiaolei Wang, Hiroki Mitoma, Daisuke Oryoji, Yuichiro Semba, Yusuke Yamauchi, Kana Yokoyama, Shotaro Kawano, Masahiro Ayano, Yasutaka Kimoto, Nobuyuki Ono, Yojiro Arinobu, Koichi Akashi, Takahiko Horiuchi, Hiroaki Niiro

## Abstract

**Background:** Systemic lupus erythematosus (SLE) is characterised by a chronic type I interferon (IFN) signature whose epigenetic basis remains incompletely defined. Histone H3 lysine 9 dimethylation (H3K9me2) is a key repressive mark that can shape DNA-methylation landscapes and antiviral responses.

**Methods:** Naïve CD4⁺ T cells were purified from 29 adults with SLE and 15 healthy donors. Genome-wide distributions of H3K9me2 and, for comparison, H3K27me3 were mapped by Cleavage Under Targets and Tagmentation (CUT&Tag). Targeted chromatin immunoprecipitation-qPCR and quantitative RT-PCR validated findings at canonical interferon-stimulated genes (ISGs). Pharmacological rescue was tested ex-vivo with ribavirin (RBV) ± the G9a inhibitor BIX01294. Therapeutic relevance was assessed in NZB/W-F1 lupus-prone mice treated with RBV (50–250 mg kg⁻¹, i.p., twice weekly for 20 weeks).

**Results:** CUT&Tag revealed a pronounced, genome-wide reduction of H3K9me2—but not H3K27me3—in SLE T cells, which segregated cases from controls by principal-component analysis. H3K9me2 loss was most evident across ISG loci and inversely correlated with ISG mRNA expression. In vitro, RBV restored H3K9me2 at ISG regions and repressed ISG transcripts; both effects were abrogated by G9a inhibition, implicating G9a-dependent dimethylation. In NZB/W-F1 mice, RBV dose-dependently reduced proteinuria, diminished renal immune-complex deposition, and normalised splenic CD4⁺ T-cell ISG expression, mirroring ex-vivo epigenetic rescue.

**Conclusions:** Loss of H3K9me2 is a selective epigenetic lesion that sustains the type I IFN signature in SLE. Pharmacological reinstatement of this mark with ribavirin reverses aberrant ISG activation and ameliorates lupus nephritis in vivo, highlighting H3K9me2 restoration as a tractable therapeutic strategy for IFN-high SLE.

## Introduction

Systemic lupus erythematosus (SLE) exemplifies a systemic autoimmune disease in which the loss of immunological self-tolerance precipitates multi-organ inflammation and progressive, cumulative tissue injury [1, 2]. Despite the availability of targeted biological therapies, a considerable subset of patients continues to experience unpredictable disease flares and the accumulation of irreversible organ damage [3].

Aberrant type I interferon (IFN) signalling constitutes one—though not the only—central pathogenic pathway in SLE. Sustained up-regulation of interferon-stimulated genes (ISGs), commonly referred to as the “IFN signature”, is detectable in approximately 60–80 % of adults and correlates with clinical activity; nevertheless, up to 40 % of patients remain signature-negative [4, 5]. The clinical efficacy of agents that block IFN-α or its receptor underscores the importance of this pathway, yet heterogeneous and often only partial responses indicate that the mechanisms sustaining chronic IFN activation—particularly during periods of apparent clinical quiescence—are still incompletely understood [2, 3, 4, 5].

Epigenetic dysregulation is increasingly recognised as a primary upstream driver rather than a mere downstream consequence of inflammation [6, 7]. Genome-wide surveys consistently demonstrate widespread DNA hypomethylation across T-, B- and myeloid-cell compartments, with differentially methylated regions clustering disproportionately at ISG loci [8–10].

Notably, naïve CD4⁺ T cells already display hypomethylation of ISG promoters before antigen encounter, implying an intrinsic cellular defect that primes these cells for exaggerated IFN responses [10, 11]. The molecular events that establish and maintain this epigenetic susceptibility remain to be elucidated.

Epigenetic regulation is orchestrated through mutually reinforcing interactions between DNA methylation and histone modifications. Repressive histone marks—H3K9me1/2/3 and H3K27me2/3—recruit DNA-methyl-transferases, thereby consolidating transcriptional silencing [12–15]. Conversely, activating modifications such as H3K4me2/3, H3K79 methylation, and deposition of the histone variant H2A.Z oppose this recruitment and shield selected genomic domains from methylation, establishing context- and lineage-specific transcriptional programmes [12, 13].

In systemic lupus erythematosus (SLE) this epigenetic balance is disrupted. Genome-wide hypo-acetylation of histones H3 and H4 in patient-derived CD4⁺ T and B cells is accompanied by aberrant—largely increased—accumulation of H3K27me3 at loci implicated in disease pathogenesis [1, 6, 16]. The extent to which such histone alterations interact with DNA methylation to modulate interferon-stimulated gene (ISG) expression remains unclear.

We therefore centred our investigation on histone H3 lysine 9 dimethylation (H3K9me2), a modification essential for the establishment and maintenance of heterochromatin [17, 18]. H3K9me2 is dynamically regulated and often heralds subsequent changes in DNA methylation [13, 18]. Recent data indicate that high H3K9me2 occupancy at interferon-related loci is associated with dampened antiviral responses [19–21]. We hypothesised that diminished H3K9me2 might preferentially release IFN-regulated genes from repression, thereby sustaining the type I IFN signature that typifies SLE [4, 5].

To test this hypothesis we mapped H3K9me2 and, for comparison, the broader repressive mark H3K27me3 across the genome of naïve CD4⁺ T lymphocytes isolated from patients with SLE and demographically matched healthy donors [8, 21]. Employing the highly sensitive Cleavage Under Targets and Tagmentation (CUT&Tag) technique on antigen-inexperienced cells enabled us to capture primary epigenetic abnormalities while minimising confounding by activation state, chronic inflammation, or therapy [21]. Given the clinical tractability of reversible chromatin marks, we further evaluated whether pharmacological restoration of H3K9me2 could normalise aberrant ISG expression [22, 23].

## Materials and methods

### Study participants

Peripheral venous blood samples were drawn from 29 individuals with systemic lupus erythematosus (SLE) and 15 healthy volunteers attending Kyushu University Hospital or Kyushu University Beppu Hospital (Japan). Participant demographics and clinical profiles are outlined in Supplementary Table 1. The study was conducted in accordance with the Declaration of Helsinki and received approval from the Ethics Committee/Institutional Review Board of Kyushu University Hospital (approval No. 29-544). Written informed consent was obtained from every participant prior to enrolment. Clinical information— including disease manifestations, laboratory data, and current as well as previous treatments—was abstracted from the patients’ medical records.

### Chromatin immunoprecipitation (ChIP) and ChIP-qPCR

A total of 5 × 10⁵ cells were cross-linked in 1 % formaldehyde for 10 min at room temperature and the reaction was terminated with stop solution (Low-Cell ChIP-seq Kit, Active Motif #53084). Pellets were lysed in ChIP buffer and sonicated for 10 min at 25 % amplitude. Pre-clearing was performed for 6 h at 4 °C with a 1:1 mixture of magnetic Protein A and Protein G beads (Thermo Fisher Scientific). Chromatin was immunoprecipitated overnight at 4 °C with 1 µg anti-H3K9me2 (Abcam #1220) or anti-H3K27me3 (Diagenode C15410069). Beads were washed twice each with low-salt wash buffer and high-salt wash buffer. DNA–protein complexes were eluted in 100 µL 1 % SDS/0.1 M NaHCO₃ at 65 °C for 1 h, followed by reversal of cross-links with 5 µL 5 M NaCl and 2 µL Proteinase K at 65 °C for 6 h. DNA was purified by phenol : chloroform : isoamyl alcohol extraction (25 : 24 : 1). Quantitative PCR was carried out with SYBR Green chemistry; primer sequences are listed in Supplementary Table 3. Because H3K9me2 is characteristically depleted at transcription-start sites but forms broad enrichment domains across gene bodies and other heterochromatic regions, primer pairs for ChIP-qPCR were placed within representative intragenic regions of each target gene rather than in the promoter. Locating amplicons in these gene-body domains maximises signal-to-noise and conforms to the canonical genomic distribution of H3K9me2 in mammalian cells [17, 19]. Enrichment values (% input) obtained from these gene-body amplicons were normalised to an H3K9me2-depleted intergenic region that served as a negative control. Enrichment was calculated as % input = 100 × 2^(Ct_input – Ct_IP).

### Cleavage under targets and tagmentation (CUT&Tag)

Naïve CD4⁺ T cells from patients with SLE or healthy donors were processed with the CUT&Tag-IT™ Assay Kit (Active Motif #53160) according to the manufacturer’s instructions. Following antibody binding, chromatin was fragmented with A-Tn5 transposase, adapters were incorporated, and libraries were prepared. Library quality was assessed on an Agilent 4200 TapeStation and sequenced on an Illumina NextSeq 550 (paired-end, × 38 bp). Approximately 8 million reads per sample were generated and aligned to the human genome (hg38).

### Sequencing data processing

Raw paired-end reads were first inspected with FastQC to evaluate overall sequence quality. Adapter contamination and low-quality bases were removed with Trimmomatic, and the filtered reads were aligned to the human reference genome (hg38) using Bowtie2. Duplicate alignments were identified and excluded with Picard MarkDuplicates. Enrichment peaks were called with SICER, after which peak lists from all samples were merged with BEDtools to construct a unified peak catalogue. Fragment counts within these regions were obtained with featureCounts, normalised by the trimmed mean of M-values method and subjected to differential analysis in edgeR; peaks with |log₂ fold change| > 1 and a false-discovery rate < 0.05 were deemed significant. Peaks were then annotated relative to transcription start sites into eight categories (1–5 kb upstream, promoter, 5′UTR, exon, intron, 3′UTR, exon– intron boundary and intergenic). Genome-wide aggregation plots and heatmaps were generated with deepTools, and motif enrichment—both known and de novo—was assessed with HOMER.

### Animals

The animal study was approved by The Ethic Committee of Kyushu University (approval no. A19-345-1). Female New-Zealand Black × New-Zealand White F1 (NZB/W-F1) mice, which spontaneously develop lupus-like disease, and age-matched female BALB/c mice were purchased from Japan SLC and Jackson Laboratory, respectively. All animals were maintained under specific-pathogen-free conditions with a 12-h light/dark cycle and free access to standard chow and water.

### Experimental protocol for the murine SLE model

At 16 weeks of age, NZB/W-F1 (n = 40) and BALB/c (n = 8) mice were randomised into six groups (8 mice per group; 4 mice per cage). NZB/W-F1 groups received intraperitoneal injections of ribavirin (RBV) at 250, 150 or 50 mg kg⁻¹, or phosphate-buffered saline (PBS) twice weekly for 20 weeks; BALB/c controls received PBS. Urine was collected every weeks in metabolic cages and proteinuria assessed semi-quantitatively with test strips (EIKEN Chemical E-UR97). Scores were defined as mild (30–100 mg dL⁻¹), moderate (100– 300 mg dL⁻¹) or severe (> 300 mg dL⁻¹). At 9 months of age, splenocytes were harvested; red blood cells were lysed with eBioscience 1 × RBC Lysis Buffer (Thermo Fisher Scientific), and CD4⁺ T cells were purified by negative selection (EasySep™ Mouse CD4⁺ T Cell Isolation Kit, STEMCELL Technologies #19852).

## Results

### H3K9me2, but not H3K27me3, delineates naïve CD4⁺ T cells from patients with SLE

To define the chromatin landscape that separates systemic lupus erythematosus (SLE) from health, we profiled naïve CD4⁺ T cells obtained from individuals with SLE and age- and sex-matched healthy donors using Cleavage Under Targets and Tagmentation (CUT&Tag) sequencing [21]. CUT&Tag was selected because it generates high-resolution maps from the limited cell numbers typically available in clinical samples. We focused on two transcriptionally repressive histone modifications—H3K9me2 and H3K27me3—both mechanistically linked to DNA methylation [12, 13]. Naïve CD4⁺ T cells (CD3⁺CD4⁺CD45RA⁺) were isolated at >98 % purity (Supplementary Fig. 1).

Genome-wide heat-maps revealed a pronounced reduction of H3K9me2 signal from the transcription start site (TSS) to the transcription end site (TES) in SLE samples relative to controls (Fig. 1A). In contrast, global H3K27me3 distribution was largely indistinguishable between the two groups (Supplementary Fig. 2A). Principal-component analysis and hierarchical clustering based on H3K9me2 abundance segregated patient and control samples into non-overlapping clusters (Fig. 1B, C), whereas H3K27me3 profiles failed to do so (Supplementary Fig. 2B, C). These data indicate a selective perturbation of H3K9me2 in SLE.

**Figure 1.**
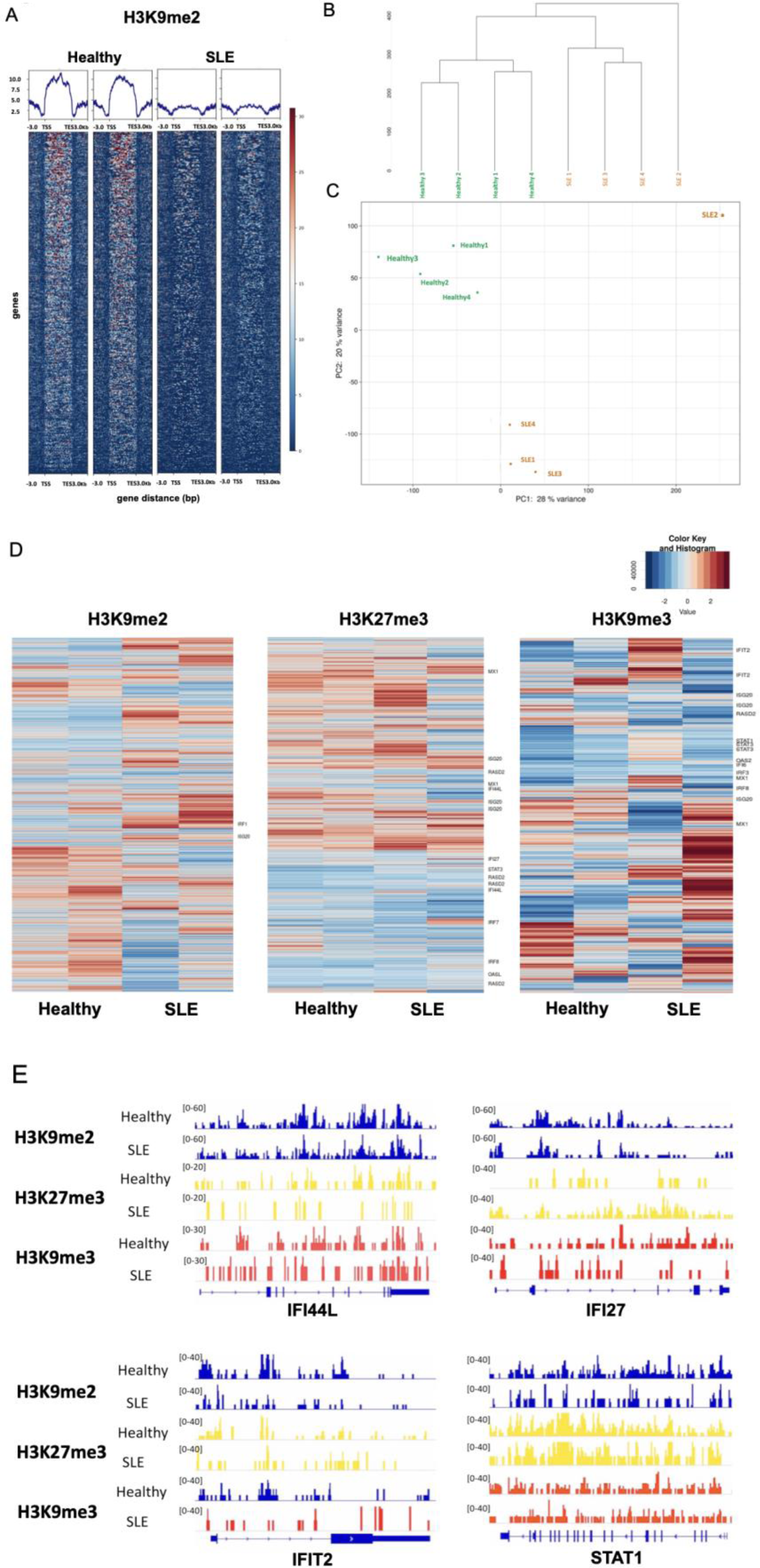
Genome-wide H3K9me2 depletion distinguishes SLE naïve CD4⁺ T cells from healthy controls. (A) Heatmap of H3K9me2 signal intensity (log₂[normalized counts + 1]) from −3 kb upstream of the TSS to the TES. Color scale: dark blue = low (0), dark red = high (8). (B) Hierarchical clustering of samples. (C) PCA plot (PC1 = 28 %, PC2 = 20 % variance). Each point = one donor (Healthy n = 4, SLE n = 5; one healthy sample failed QC and is omitted from panels A–C but included in statistical aggregates; see Methods). (D) Gene-level heatmaps of H3K9me2, H3K27me3, and H3K9me3 for representative ISGs. (E) IGV tracks showing H3K9me2, H3K27me3, and H3K9me3 at IFI44L, IFI27, IFIT2, and STAT1 loci (representative donor from each group).

A heat-map of differentially enriched regions underscored this divergence (Fig. 1D). Because interferon-stimulated genes (ISGs) generally reside in chromatin with low activity scores, H3K9me2 peaks were scarcely detected over ISG loci. In contrast, H3K9me3 and H3K27me3 signals over prototypic ISGs—MX1, ISG20, IRF8, RASD2 and STAT3—were comparable between patients and controls. Notably, ISG loci such as IFI27, IFI44L, IFIT2 and STAT1 exhibited a marked depletion of H3K9me2 in SLE. Integrative Genomics Viewer tracks confirmed higher H3K9me2 occupancy at these loci in healthy donors, consistent with physiological ISG silencing (Fig. 1E). Levels of H3K9me3 and H3K27me3 at the same loci remained unchanged (Fig. 1E). Collectively, these observations identify loss of H3K9me2— not H3K27me3—as a distinctive epigenetic hallmark that may underlie aberrant ISG activation in SLE.

### Reduced H3K9me2 at interferon-stimulated gene (ISG) loci in naïve CD4⁺ T cells from patients with systemic lupus erythematosus

Building on our genome-wide survey, we asked whether the diminution of H3K9 dimethylation (H3K9me2) at ISG loci mechanistically contributes to the well-known ISG over-expression in systemic lupus erythematosus (SLE). Quantitative RT-PCR confirmed a pronounced up-regulation of the canonical ISGs *MX1*, *IFI27*, and *IFI44L* in SLE samples (Fig. 2A), reiterating the persistent type I interferon signature that typifies the disease.

**Figure 2.**
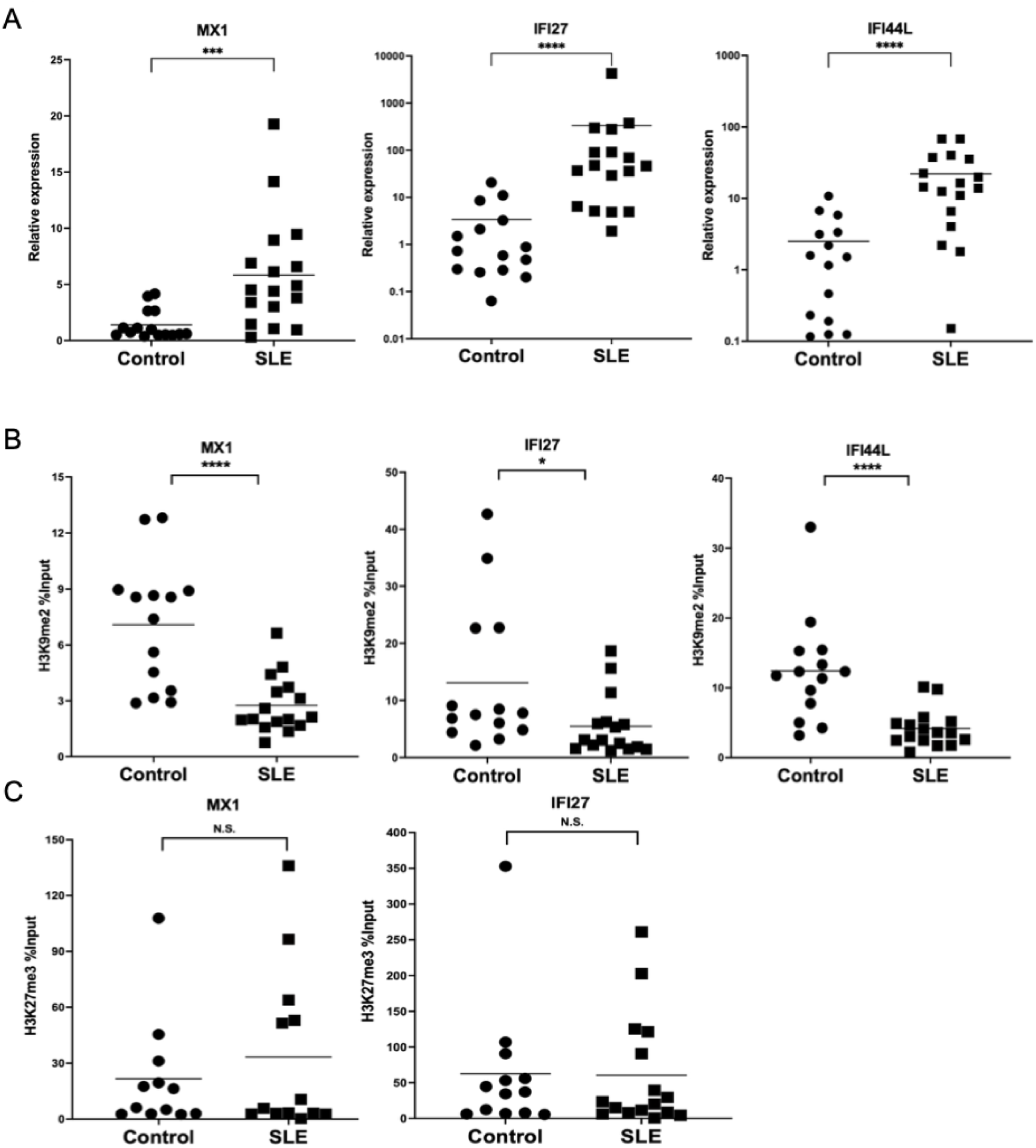
Interferon-stimulated genes (ISGs) are up-regulated and lose H3K9me2 in SLE naïve CD4⁺ T cells. (A) ISG mRNA expression; each dot = one donor (Healthy n = 15, SLE n = 29); medians shown. (B) H3K9me2 ChIP-qPCR (% input) at selected ISGs (same donors as A). (C) H3K27me3 ChIP-qPCR. Statistics: two-tailed Mann–Whitney U test; \**P* < 0.05, \*\**P* < 0.01, \*\*\**P* < 0.001, \*\*\*\**P* < 0.0001; ns = not significant. Data plotted on log₁₀ scale where necessary to display zero values.

To address the role of repressive histone modifications, we performed chromatin immunoprecipitation followed by quantitative PCR (ChIP-qPCR) for H3K9me2 and the alternative repressive mark H3K27me3 at the promoters of these genes. In agreement with the genome-scale data set, H3K9me2 occupancy was markedly reduced in SLE cells relative to healthy controls (Fig. 2B), and the degree of loss inversely correlated with transcript abundance. By contrast, H3K27me3 enrichment did not differ between the two groups (Fig. 2C), indicating that the impact of autoimmunity on repressive chromatin is mark-specific.

Collectively, these observations delineate a focused epigenetic lesion—failure to deposit H3K9me2 at ISG regions—that facilitates pathological ISG de-repression in SLE. The selectivity of this defect implies a functional impairment of the enzymatic machinery responsible for H3K9 dimethylation, thereby sustaining a maladaptive, interferon-driven transcriptional programme in naïve CD4⁺ T cells.

### Ribavirin (RBV) re-establishes H3K9me2 at interferon-stimulated gene loci and mitigates the type I IFN signature in naïve CD4⁺ T cells

We next asked whether the epigenetic defect could be pharmacologically corrected. Ribavirin (RBV), an approved antiviral reported to promote H3K9me2 deposition at ISG promoters in other settings [22], was therefore examined. Naïve CD4⁺ T cells isolated from patients with SLE were stimulated via CD3/CD28 and incubated with RBV for 72 h. Quantitative RT-PCR showed a marked reduction in the expression of representative ISGs (MX1, IFI27, IFI44L) relative to vehicle-treated cells (Fig. 3A).

**Figure 3.**
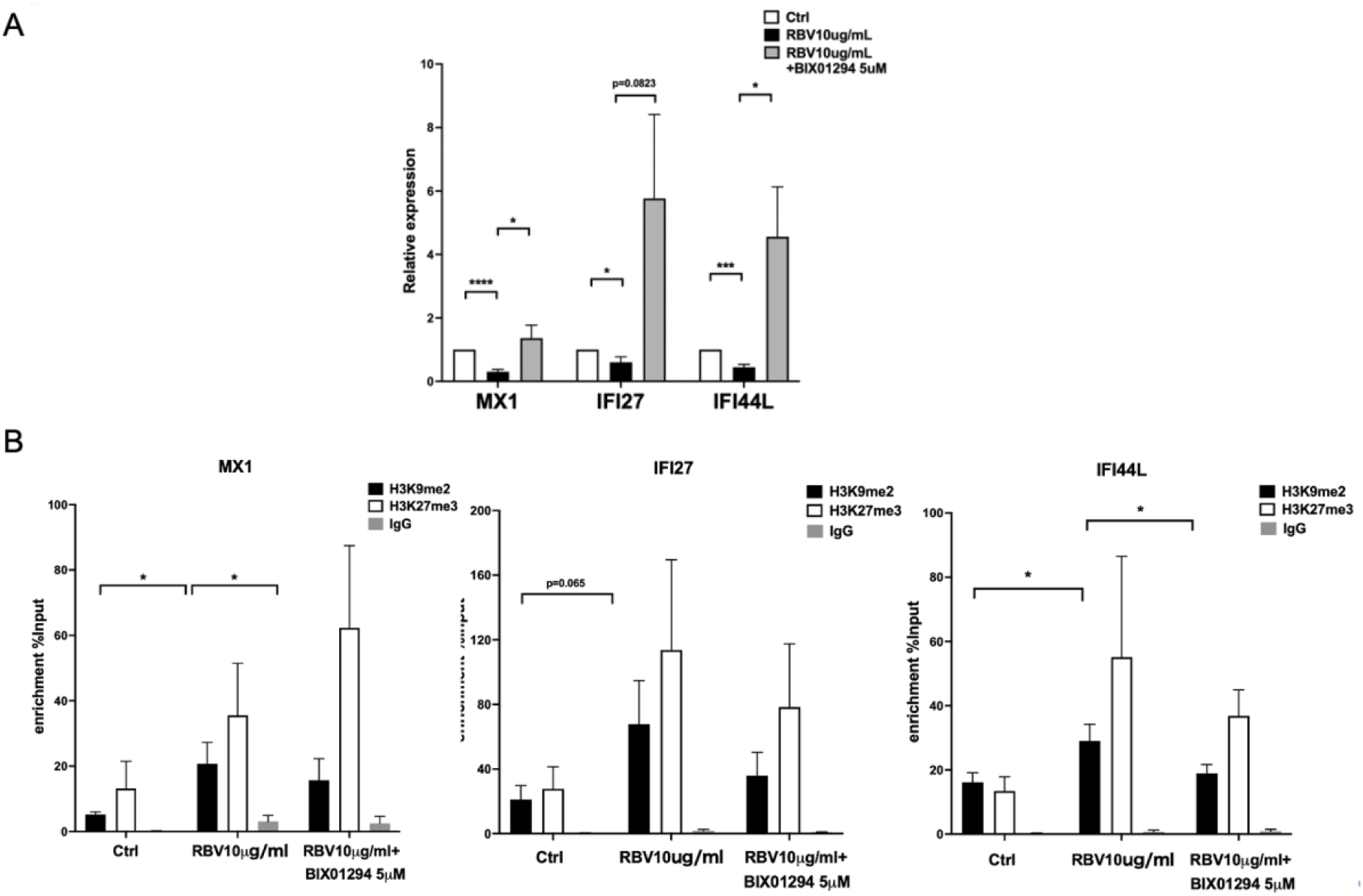
Ribavirin (RBV) restores H3K9me2 and represses ISGs in vitro. Naïve CD4⁺ T cells from three independent SLE patients were activated ± RBV (10 μg mL⁻¹) for 72 h with or without the G9a inhibitor BIX01294 (5 μM). (A) Relative ISG mRNA expression (mean ± SEM, n = 3). (B) ChIP-qPCR (% input) for H3K9me2 and H3K27me3 at ISG regions (mean ± SEM, n = 3). One-way ANOVA with Tukey post-hoc; exact P values shown where space permits; asterisks as in Fig 2.

To determine whether this repression required G9a-dependent H3K9me2, cells were co-treated with the selective G9a inhibitor BIX01294. BIX01294 completely reversed RBV-mediated ISG silencing (Fig. 3A), identifying G9a as an essential effector of the rescue. Consistently, ChIP-qPCR demonstrated that RBV significantly augmented H3K9me2 at the same genes, whereas this enrichment was lost upon G9a inhibition (Fig. 3B). RBV left H3K27me3 unchanged, underscoring the specificity of the H3K9me2 pathway (Fig. 3B).

Genome-wide CUT&Tag confirmed these locus-specific observations: RBV increased H3K9me2 occupancy at canonical ISGs—including IFI44L, IRF8 and STAT3—(Fig. 4A, 4C). Unsupervised clustering and principal-component analysis segregated RBV-treated samples from controls on the basis of their restored H3K9me2 landscape, whereas H3K27me3 profiles remained largely unaltered (Fig. 4B; online Supplementary Fig. 2C). Collectively, these data indicate that RBV reinstates the repressive H3K9me2 mark at ISG regions, thereby dampening the aberrant interferon programme characteristic of SLE.

**Figure 4.**
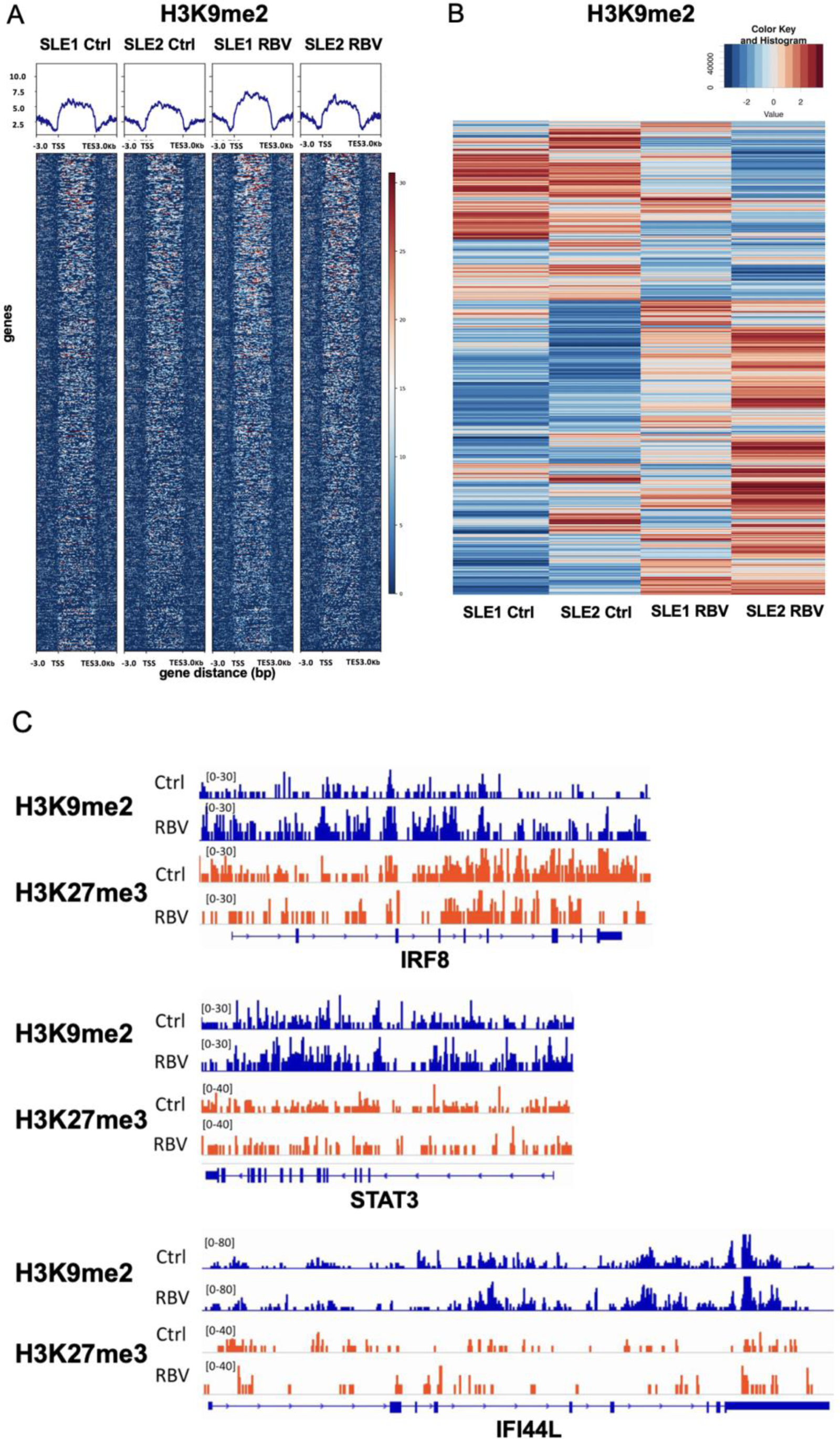
Genome-wide epigenetic rescue by RBV. CUT&Tag on naïve CD4⁺ T cells from two SLE donors (SLE1, SLE2) after 72 h culture ± RBV (10 μg mL⁻¹). (A) Heatmap of H3K9me2 signal (color scale as Fig 1A). (B) Differential H3K9me2 cluster heatmap (Z-score). (C) Browser views of IRF8, STAT3, and IFI44L loci (representative donor). Data are representative of two biological replicates; complete dataset deposited in GEO (accession GSE274177).

### Ribavirin mitigates lupus nephritis in NZB/W F1 mice

To determine whether restoration of H3K9me2 by ribavirin (RBV) confers clinically meaningful benefit, we employed NZB/W F1 mice, the canonical model of spontaneous SLE-like glomerulonephritis. Previous work documented partial protection by RBV in this strain [25, 26]. Beginning at 3 months of age, animals received intraperitoneal injections twice weekly until 9 months of age with either phosphate-buffered saline (PBS), cyclophosphamide (CYC; positive control), or RBV at 50, 150, or 250 mg kg⁻¹. Serial quantification of proteinuria—a cardinal marker of lupus nephritis—revealed a graded decline across the RBV groups; the 250 mg kg⁻¹ dose achieved proteinuria scores comparable to CYC (Fig. 5A).

**Figure 5.**
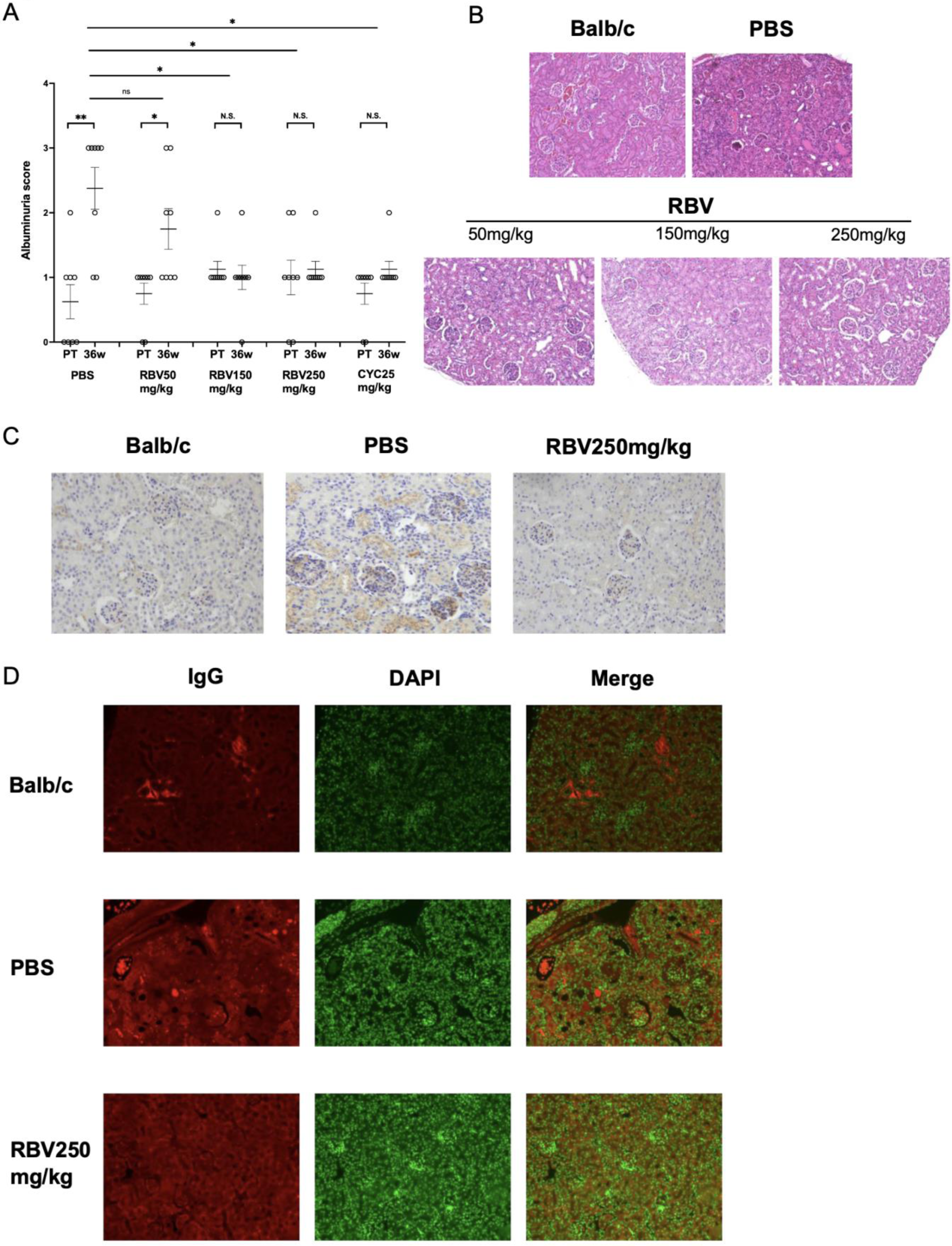
RBV ameliorates lupus nephritis in NZB/W F1 mice. Female NZB/W F1 mice were treated twice weekly with RBV (50, 150, 250 mg kg⁻¹), cyclophosphamide (CYC 25 mg kg⁻¹) or PBS from 16 to 36 weeks of age (n = 8 per group). (A) Albuminuria score (median ± IQR). Horizontal bars = Kruskal–Wallis with Dunn’s post-hoc vs. PBS; *P < 0.05, **P < 0.01; ns = not significant. (B) H&E kidney sections (representative). (C) C3 immunohistochemistry. (D) IgG immunofluorescence (left, red), DAPI (middle, green), merged (right). Scale bars = 100 μm.

**Figure 6.**
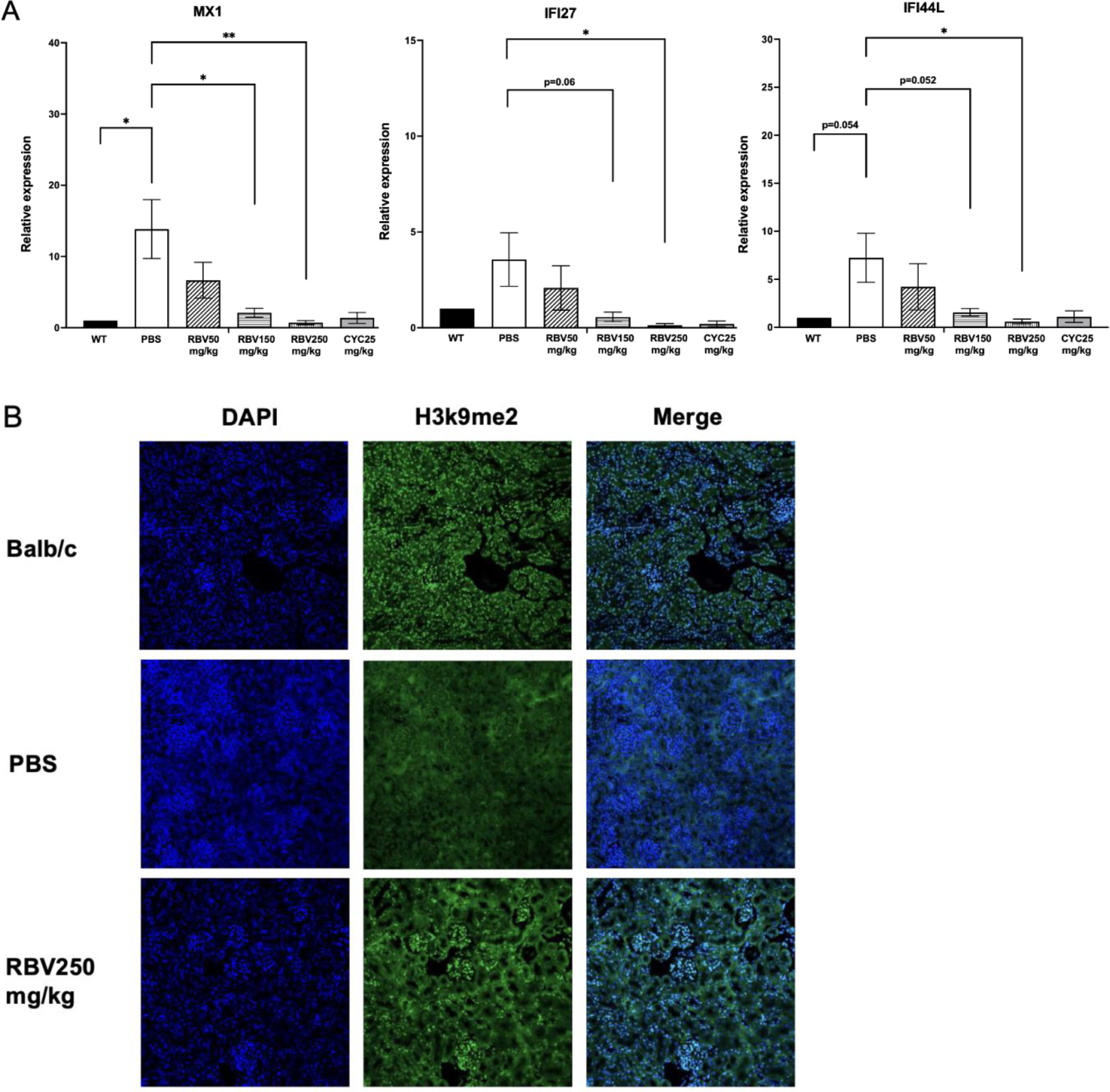
RBV normalizes ISG expression and H3K9me2 in vivo. (A) Splenic CD4⁺ T-cell ISG mRNA (mean ± SD, n = 8). One-way ANOVA with Tukey; *P < 0.05, **P < 0.01; dotted lines indicate comparisons where P = 0.054–0.06 (trend). (B) Kidney immunofluorescence for H3K9me2 (green) with DAPI counterstain (blue); representative fields, scale 50 μm.

Renal histopathology supported these functional data. Haematoxylin–eosin sections showed diminished interstitial inflammatory infiltrates, fewer karyorrhectic nuclei, and reduced tubular cast formation in RBV-treated mice relative to PBS controls (Fig. 5B). Immunohistochemistry for complement component 3 demonstrated attenuated immune-complex deposition (Fig. 5C), while DAPI counter-staining confirmed decreased leukocyte influx. Consistently, immunofluorescence revealed lower IgG accumulation in glomeruli and tubules (Fig. 5D).

Collectively, these observations indicate that epigenetic correction by RBV translates into substantive in-vivo renoprotection in NZB/W F1 lupus nephritis.

## Discussion

Here, we report that naïve CD4⁺ T cells from patients with systemic lupus erythematosus (SLE) exhibit a pronounced loss of H3K9 dimethylation (H3K9me2) across interferon-stimulated gene (ISG) domains. This targeted erosion of a repressive histone mark offers mechanistic insight into the entrenchment of the canonical type I interferon signature in SLE. Our data indicate that the defect is unlikely to be a mere downstream consequence of chronic inflammation; rather, it appears early—paralleling the first descriptions of epigenetic disturbance in lupus T cells [27]—and helps sustain aberrant IFN-driven transcription. Consistent with this concept, H3K9me2 occupancy was markedly diminished at several prototypical ISGs in SLE samples, whereas the distribution of the alternative repressive mark H3K27me3 remained largely comparable to that in healthy donors. The marked selectivity argues for malfunction of the enzymatic machinery responsible for depositing H3K9me2, including EHMT1/EHMT2 and related lysine-methyltransferases [28, 18].

Whether loss of H3K9me2 precedes, or instead follows, alterations in DNA methylation cannot be resolved from the present data set. Nevertheless, because H3K9me2 frequently functions as an early chromatin cue that guides subsequent DNA methylation [12, 13], persistent depletion of this mark could precipitate stable changes in the DNA methylome. Such a cascade accords with the well-documented crosstalk between histone modifications and DNA methylation mediated by MBD “reader” proteins [29] and is compatible with longitudinal evidence that epigenetic patterns remain durably altered in lupus neutrophils [33].

By showing that ribavirin (RBV) reinstates H3K9me2 at ISG regions and concomitantly alleviates the cellular and clinical manifestations of SLE, we underscore the therapeutic promise of targeting histone methylation. In our murine model, administration of RBV at 250 mg kg⁻¹ reduced proteinuria by approximately 50 %—a magnitude comparable with that achieved by cyclophosphamide—and markedly diminished glomerular immune-complex deposition, echoing earlier reports in NZB/W F₁ mice [25, 26]. Although G9a/EHMT2 is generally regarded as the predominant H3K9 dimethyl-transferase, the steady-state abundance of H3K9me2 in SLE represents the net balance between methyltransferase and demethylase activities. Members of the KDM3 (JMJD1) family, which remove mono- and dimethyl groups from H3K9, are up-regulated under oxidative stress in lupus T cells [30] and may accentuate H3K9me2 loss. Histone demethylases in the KDM4 family (e.g. KDM4D/JMJD2d) likewise fine-tune antiviral responses [31]. We demonstrate that RBV partially restores the H3K9me2 mark and represses aberrant ISG expression, thereby interrupting the pathological loop in which sustained IFN signalling recruits demethylases or impedes H3K9 methyltransferases [22].

An unresolved issue is whether simultaneous blockade of KDM3 demethylases would amplify the therapeutic benefits observed with RBV alone. Systematic evaluation of combination regimens—for example, co-administration of RBV with highly selective KDM3 inhibitors—could restore physiological transcriptional control more effectively and deliver additive or even synergistic efficacy relative to single-agent therapy. Such strategies, however, must be preceded by comprehensive toxicological profiling because KDM3 family members participate in numerous epigenetic networks well beyond repression of ISGs [23].

Our data likewise underscore the need to interrogate additional immune compartments that drive SLE. Abnormal type I IFN signalling is also documented in B cells, dendritic cells and myeloid subsets [32, 24, 20], each of which may harbour shared—or cell-specific— dependencies on balanced H3K9me2 dynamics. Defining the repertoires of G9a- and KDM3-controlled loci in these populations will determine whether RBV operates through analogous mechanisms outside the T-cell compartment. Concomitantly, elucidating how H3K9me2 crosstalks with other repressive histone marks should refine our understanding of chromatin-level regulation in SLE pathogenesis.

In the NZB/W F₁ lupus model, prophylactic administration of RBV before nephritis onset consistently postponed the appearance of clinical disease [25, 26], indicating that timely epigenomic modulation can impede establishment of a pathogenic chromatin landscape. Genome-wide association studies and integrative multi-omics analyses demonstrate that many SLE risk alleles perturb immune tolerance during the clinically silent phase [34]—a period marked by low-level autoantibody production [33]. We therefore propose that introducing RBV in genetically predisposed individuals could forestall the locking-in of aberrant histone marks and, in turn, pre-empt progression to overt autoimmunity. A composite strategy integrating genetic risk stratification, ISG profiling and epigenetic read-outs may accurately identify candidates for such prophylactic therapy.

RBV may also serve as a remission-maintenance agent. Contemporary induction regimens, centred on glucocorticoids and cytotoxic immunosuppressants, achieve disease control but impose substantial cumulative toxicity. By stabilising repressive chromatin states, RBV could allow earlier tapering of conventional immunosuppression and thus mitigate adverse events. Nevertheless, the pronounced clinical, serological and genetic heterogeneity of SLE makes a uniform response improbable. Carefully designed biomarker-guided trials that stratify participants by baseline IFN load, histone-methylation pattern and disease activity are required to delineate the patient subsets in which RBV will be most efficacious.

Translating RBV-mediated epigenetic reprogramming to routine care still presents several hurdles. Dose-finding studies must balance sufficient histone re-methylation against the risk of cumulative toxicity, while long-term follow-up is necessary to confirm persistence of benefit and to detect delayed disease reactivation. Equally important is rigorous pharmacovigilance: RBV was purpose-built for antiviral therapy and its safety profile in an autoimmune milieu remains incompletely characterised. Early, small-scale human trials reported limited efficacy and highlighted anaemia as a dose-limiting toxicity [35]. Although the present work is limited to the NZB/W F₁ model, these data reinforce the broader principle that targeted remodelling of the immune epigenome can curtail disease propagation and lessen exposure to conventional immunosuppression. By identifying depletion of H3K9me2 as a tractable lesion, we provide a mechanistic rationale for early or adjunctive RBV therapy to disrupt the interferon-amplifying loop that sustains lupus activity. Future studies integrating multi-omics and rational drug combinations should solidify the translational pathway for epigenetic intervention in SLE.

## Data Availability

The raw sequencing data generated in this study have been deposited in the Gene Expression Omnibus (GEO) database under accession code GSE274177.

Conceptualisation: Qiaolei Wang, Daisuke Oryoji, Hiroki Mitoma

Methodology: Qiaolei Wang, Daisuke Oryoji, Yuichiro Semba

Investigation: Qiaolei Wang, Yusuke Yamauchi, Kana Yokoyama

Writing ‒ original draft: Daisuke Oryoji, Qiaolei Wang

Writing ‒ review & editing: all authors

Funding acquisition: Hiroki Mitoma, Daisuke Oryoji

Supervision: Hiroaki Niiro, Takahiko Horiuchi

## Data availability

Sequencing data have been deposited in GEO under accession number GSE274177.

## Ethics approval and consent Human study

Kyushu University IRB #29-544 (written informed consent obtained). Animal study: Kyushu University Animal Ethics Committee #A19-345-1.

## Supplementary Figure Legends

**Supplementary Figure 1.**
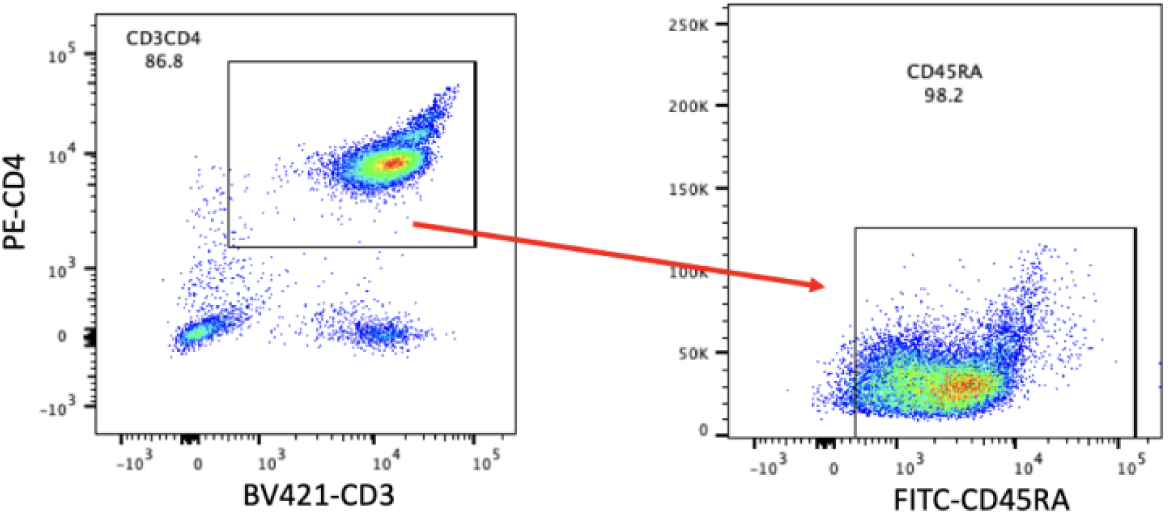
Purity of sorted naïve CD4⁺ T cells. Representative flow-cytometry gating strategy; overall purity > 98 % (n = 8 runs).

**Supplementary Figure 2.**
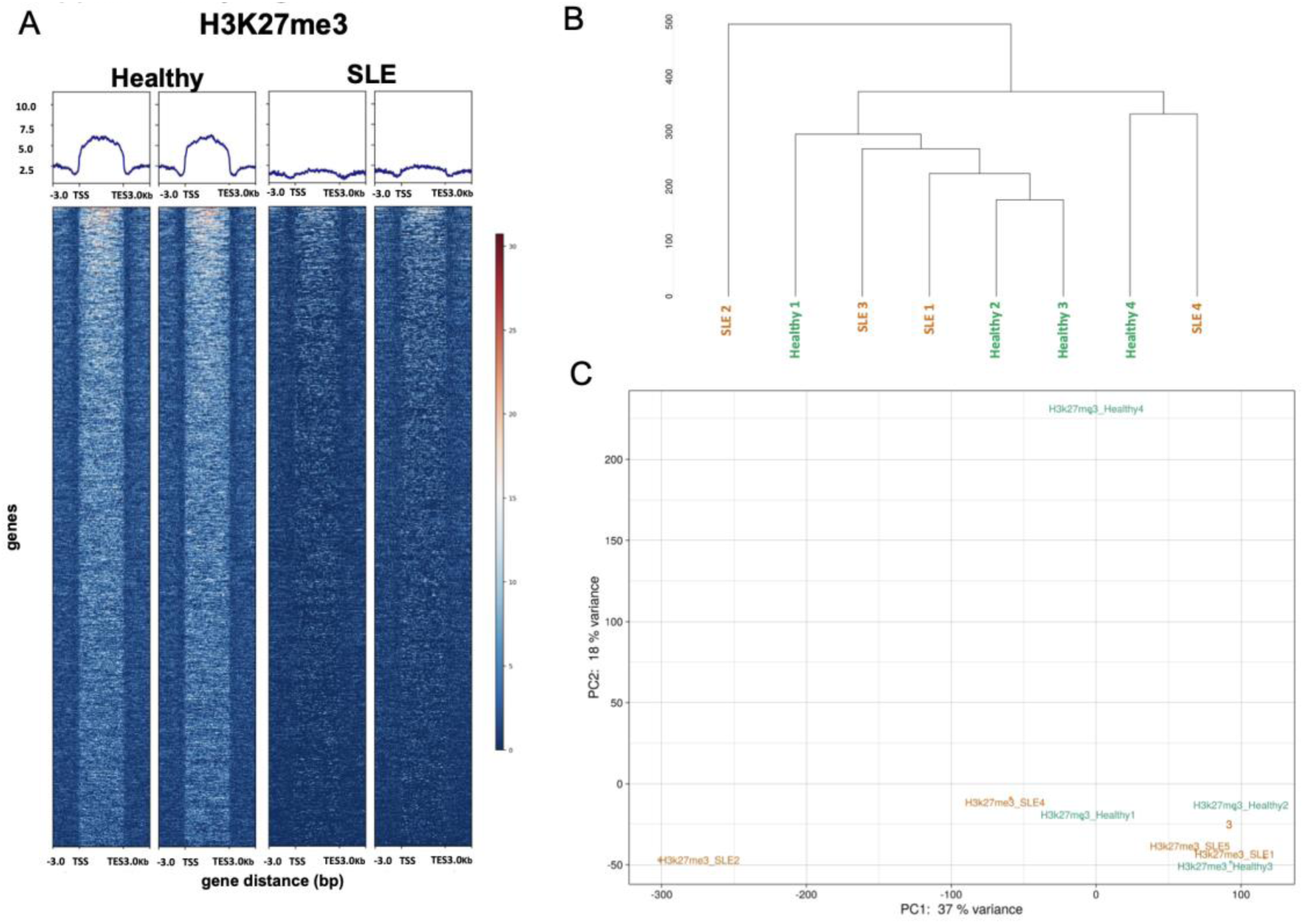
H3K27me3 landscape in SLE naïve CD4⁺ T cells. Heatmap, clustering, and PCA as in Fig 1 but for H3K27me3 (Healthy n = 4, SLE n = 5).

**Supplementary Figure 3.**
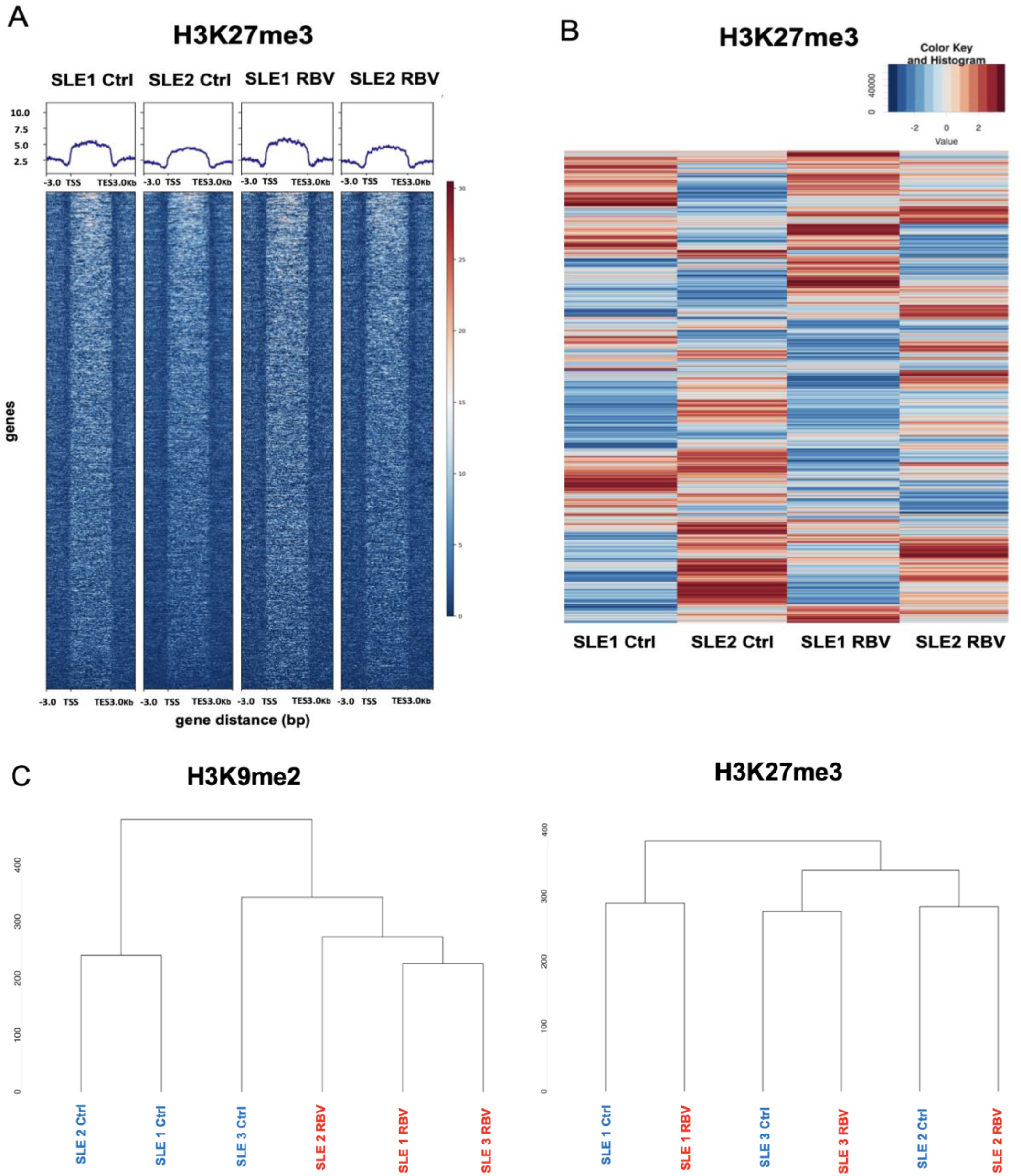
H3K27me3 is not restored by RBV. CUT&Tag on SLE donors 1 & 2 ± RBV (10 μg mL⁻¹). Heatmap, differential cluster, and hierarchical dendrogram for H3K27me3.

## Supplementary Tables

**Supplementary Table 1.** Clinical characteristics of SLE cohort (n = 29).

| Characteristics |  |
| --- | --- |
| Age, mean, years | 41.1±15.1 |
| Females, n (%) | 29 (100) |
| Diseases duration, median, years | 8 ± 9.4[5-17] |
| Increased anti-dsDNA antibodies, n (%) | 12 (44.4) |
| Low complement, n (%) | 19 (65.5) |
| C3, median, mg/dl | 70 ± 17.7[61-80] |
| C4, median, mg/dl | 15 ± 7.7[7-18] |
| Corticosteroid use, n(%) | 26 (89.7) |
| HCQ use, n(%) | 19 (65.5) |
| Immunosuppressive agent use, n(%) | 23 (79.3) |

HCQ, hydroxychloroquine
Variables are described using mean ±S.D. and median [IQR]

**Supplementary Table 2.** Interferon-stimulated gene (ISG) panel used for methylation analysis.

| Gene Symbol |  |
| --- | --- |
| MX1 | OASL1 |
| MX2 | OASL2 |
| IFI6 | OASL3 |
| IFI27 | OASL |
| IFI44L | RASD2 |
| IFIT1 | STAT1 |
| IFIT2 | STAT2 |
| IFITM2 | STAT3 |
| IRF1 |  |
| IRF3 |  |
| IRF5 |  |
| IRF7 |  |
| IRF8 |  |
| ISG15 |  |
| ISG20 |  |

**Supplementary Table 3.**
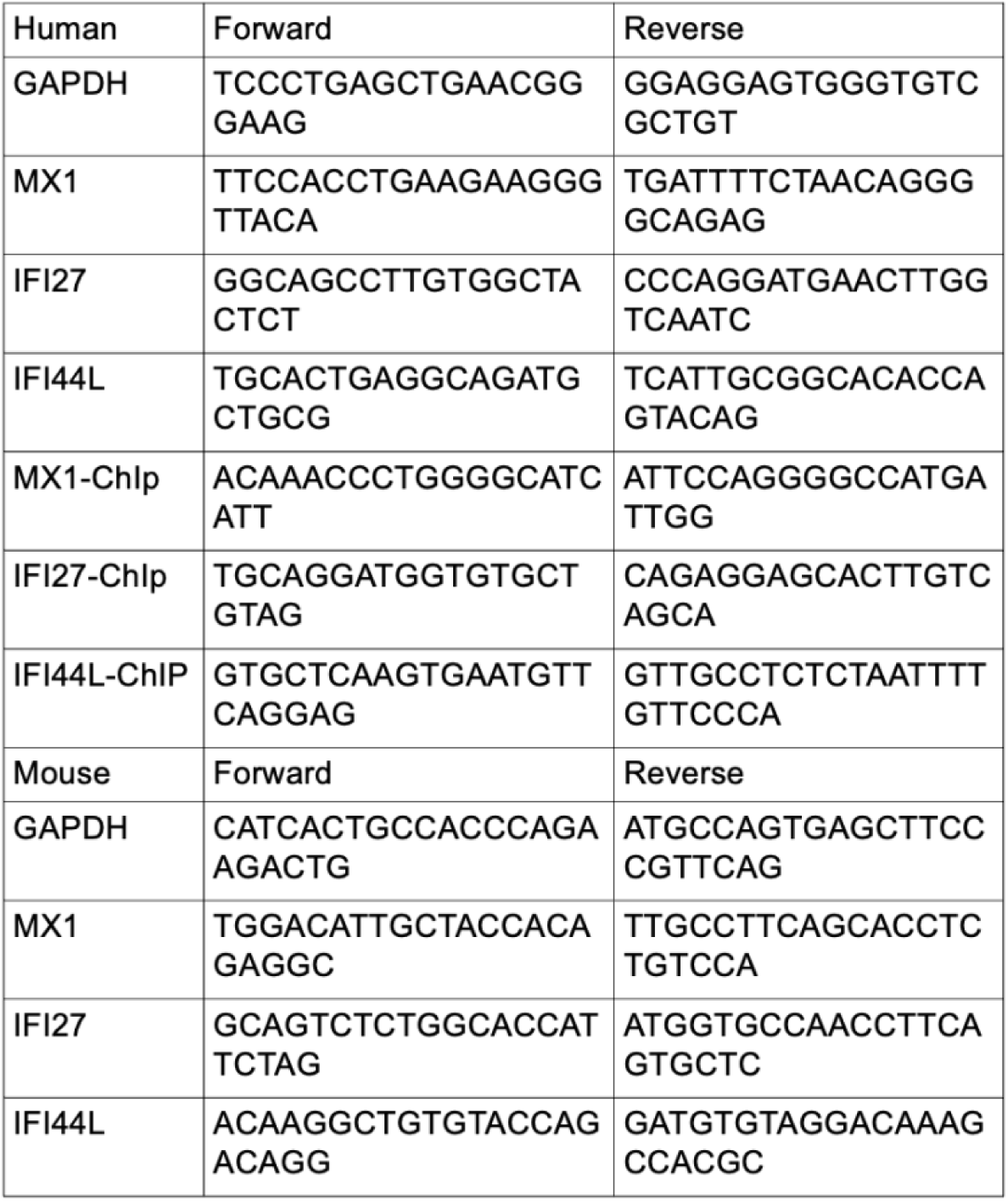
Primer sequences for qRT-PCR and ChIP-qPCR (human and mouse).

